# The Protocol for: Do peers enhance alcohol use outcomes in group motivational interviewing? A translational clinical trial investigating brain synchrony in underage emerging adults during a fMRI hyperscanning task and its association with 12 month alcohol use reductions

**DOI:** 10.64898/2025.12.10.25341908

**Authors:** Sarah W. Feldstein Ewing, Genevieve F. Dash, Karen A. Hudson, Benedict Hyun, Manshu Yang, Linnea Stahl, Kristine Rømer Thomsen, Ana Paula Gonçalves Donate, Francesca M. Filbey

## Abstract

**Background and Aims:** Alcohol is the top substance used by US emerging adults under legal drinking age (underage emerging adults; U-EA; ages 18-19). U-EA are unlikely to seek or receive indicated alcohol interventions. It is imperative to find brief, effective interventions to reduce U-EA drinking. Developmental neuroscience reflects that peers hold higher neural salience during adolescence, as evidenced by differential neural response with real and/or simulated peers, even when those peers were not friends, and particularly in the context of alcohol. Peers also activate positive (prosocial) neural and behavioral responses among adolescents. The role of positive, prosocial peer feedback is consequential given that the most widely-used U-EA platforms for adolescent addiction intervention are group-based interventions. We propose functional magnetic resonance imaging (fMRI) hyperscanning (tandem dyadic scanning) to evaluate U-EA brain response in the underexamined, but widely-utilized, group motivational interviewing (MI). We will examine associations between youth brain synchrony generated in an fMRI hyperscanning task and alcohol use reductions at 12 month follow-up. We hypothesize that greater neural alignment in the social cognition network during positive, prosocial peer interactions (peer-directed change talk) will be associated with greater post-intervention behavior change (lower number of past month drinking days) at 12 months.

**Design:** Within-subjects

**Setting:** UConn Health and UT-Dallas

**Participants:** This study will enroll n=248 U-EA (ages 18-19) with ≥1 past month binge drinking event (4/3 standard drinks per occasion for males/females) via community and campus recruitment

**Intervention and comparator:** Group MI (no comparator; within-subjects)

**Measurements:** This protocol will utilize synchronized fMRI (hyperscanning) to examine positive, prosocial peer-directed health promotive language (peer-directed change talk) directly generated during and extracted from the group MI session. Primary outcomes include fMRI-based hyperscanning metrics (BOLD synchrony in social cognition networks) and behavioral measures (past month alcohol use days measured viaTimeline Followback; TLFB) at 12 months. Secondary outcomes will evaluate the same relationship at 3- and 6-months.

**Comment:** This translational study is crucial for making meaningful gains in adolescent addiction intervention. We could find no peer-reviewed studies examining fMRI hyperscanning in an adolescent addiction intervention context. And, fMRI hyperscanning is at the forefront of interactive, dyadic youth research in socially-relevant and ecologically valid settings. In addition, due to the limited empirical evaluations of group interventions, including group MI, the use of fMRI hyperscanning is timely and offers a unique examination with high clinical impact for young people struggling with addiction.

## ADMINISTRATIVE INFORMATION

### Protocol version

Version 1.0, effective as of first enrollment 10/10/2025. No protocol changes since first enrollment.

### Roles and responsibilities

Sarah W. Feldstein Ewing, PhD: MPI overseeing the trial, project conceptualization, funding acquisition, oversight of clinical components, writing-original draft, - review and editing

Genevieve F. Dash, PhD: writing-review and editing, clinical interventionist

Karen A. Hudson, MCR: management of personnel and funding, writing-review and editing

Benedict Hyun, BS: neuroimaging task, neuroimaging data management, writing-review and editing

Manshu Yang, PhD: behavioral data analysis, writing-review and editing

Linnea Stahl, MS: project administration and coordination, writing-review and editing

Kristine Rømer Thomsen, PhD: writing-review and editing

Ana Paula Gonçalves Donate, MS: writing-review and editing

Francesca M. Filbey, PhD: MPI overseeing the trial, project conceptualization, funding acquisition, oversight of neuroimaging components, writing-original draft, - review and editing

### Name and contact information for the trial sponsor

This study is funded by the National Institute on Alcohol Abuse and Alcoholism (NIAAA; grant numbers 7R01AA030678-02 to SFE and FF). The study is sponsored by UConn Health and the University of Texas at Dallas and led by MPIs Feldstein Ewing and Filbey.

### Role of trial sponsor and funder

UConn Health, University of Texas at Dallas, and the funder have no influence on study design, collection, management, analysis and interpretation of data, writing of the report and the decision to submit for publication. MPIs Feldstein Ewing and Filbey are responsible for initiation, management, and oversight of the trial. There are no other individuals and/or groups overseeing the study.

### Composition, roles, and responsibilities of the coordinating site, steering committee, endpoint adjudication committee, data management team, and other individuals or groups overseeing the trial

Not applicable.

## OPEN SCIENCE

### Trial registration

https://ClinicalTrials.gov

ID: NCT06115252

URL: https://clinicaltrials.gov/study/NCT06115252

Date of registration: 10/30/2023

### Protocol and statistical analysis plan

In line with trial registry requirements, the trial protocol and statistical analysis plan will be available alongside results reporting on https://ClinicalTrials.gov within one year of study completion (i.e., final data collection of primary outcome).

### Data sharing

This study will submit and share data with the NIAAA Data Archive, a data repository housed within the NIMH Data Archive, and will follow the guidelines described in NOT-AA-23-002. Data will be submitted on or before the NDA submission due dates and maintained for the duration required by NIAAA Data Archive Data Sharing Plan.

### Sources of funding and other support

This work was supported by the National Institute on Alcohol Abuse and Alcoholism (NIAAA; grant numbers 7R01AA030678-02 to SFE and FF).

### Financial and other conflicts of interest

Competing interests: none declared.

### Plans to communicate trial results (dissemination policy)

Trial results will be submitted to the trial registry at https://ClinicalTrials.gov in line with registry reporting requirements. The study team will disseminate results at scientific conferences and in peer reviewed publications. MPI Feldstein Ewing conducts clinical trainings for professionals working with substance using individuals; thus, research outcomes from this project will be directly distributed to direct care providers as part of these trainings.

## INTRODUCTION

### Background and rationale

Alcohol continues to be the top substance used by underage emerging adults (U-EA) (1, 2) and 2021 data reflect that youth drinking is on the rise throughout the US (2). In addition to potential alcohol-related negative health, safety, and neurodevelopment sequalae (3–6), only a fraction (<6%) of youth engaged in hazardous drinking receive intervention programming (7). Among those who do, behavior change in this age group still remains modest (8, 9). As one example, meta-analyses examining motivational interviewing (MI) have shown that effect sizes for MI intervention outcomes are less robust for youth (d = 0.17) (10) (11) as compared with adults (d = 0.77) (12). While the use of MI is widespread, we still do not yet understand within-session mechanisms, particularly for youth and for the group MI modality (13).

Originally developed and validated as an individual-level intervention (12, 14–16), MI has been widely deployed in direct practice settings as a group-based intervention for over two decades. Yet, the empirical research on intervention outcomes with group MI, particularly among youth recipients, has lagged behind individual-level evaluations (17). Importantly, a handful of teams, including our own, have evaluated group MI across a wide range of health behaviors (including, but not limited to: alcohol and other substance use; STI/HIV risk behavior). Overall, findings largely support group MI as a modality that effectively generates behavior change in the targeted behavior (17–44). An even smaller handful of investigators have taken this work a step further by evaluating mechanisms of group MI, including within-session factors that impact group MI outcomes with youth. Collectively, these findings suggest that, among young participants, positive statements in favor of change (change talk) during group MI have been associated with significant improvements in post-intervention alcohol-related behavior (23, 27, 45). We could only find one peer-reviewed published study looking at group MI delivered in a peer-dyad format; in this study, effect sizes for alcohol use reductions were 3 times larger in the peer-dyad group MI than for the individually-delivered format (46). We found no studies looking at the impact of the peer-peer exchange on brain response.

Large-scale longitudinal studies like IMAGEN, ABCD, and NCANDA (47–49) have generated an enormous array of foundational data on the nature of the developing brain (3, 4, 50). Yet, most have examined the youth brain on an individual level, as has our own translational work to date on the neural mechanisms undergirding adolescent neural response to successful behavioral interventions (51–55). However, humans are social animals, and this is particularly true for underage emerging adults (U-EA) (56–65).

A central challenge of individual-level neuroimaging is that we can only examine one member of the peer dyad at a time (66). This is critical because youth show both enhanced neural attributions to peers, as well as modifications to behavior based on youths’ real-world and purported peer behavior (56–64), even when those peers were not friends (67–71), and particularly in the context of alcohol (72–77). Many studies have focused on the negative role of peer feedback, yet current neurodevelopmental research is increasingly indicating that peers also activate positive (prosocial) neural and behavioral responses in this age group (78–86). Yet, our capacity to examine interactive, real-world peer dyads via neuroimaging is clunky at best (66). Because most accessible and widely-used neuroimaging modalities with youth can only accommodate a single participant at a time, many studies are limited to less-ecologically valid, “proxy” social environments to estimate/approximate youth neural response in these social contexts (e.g., cyberball; images of pretend similar-aged peers; use of a confederate (76, 85, 87, 88). Some have even argued that the generalizability of these neuroimaging findings to real-world youth behavior is insufficient (66).

#### Interbrain synchrony to identify patterns of neural response in group MI

One cutting-edge solution to these conundrums is the use of hyperscanning (89). In hyperscanning, two neuroimaging platforms (e.g., two MRIs) are interconnected, allowing scientists to examine not only parallel neural response between two participants in-real time (66, 89), but, even more novel, participants’ interbrain synchronization (synchrony). Prior hyperscanning data has begun to reflect that heightened synchrony during human social interactions is linked with a range of prosocial and adaptive social behaviors, including closeness, cooperation, and other collaborative behaviors (90).

Synchrony has been examined across a number of dyad types (e.g., educator:student; teammate:teammate; employer:applicant) (91–95). In terms of neurodevelopment, existing hyperscanning studies (Total N=126; N=32 adolescent pairs; N=31 adult pairs) reflect significant developmental differences between adolescents and adults in synchrony, with adolescent dyads showing significantly higher synchrony than adults in relevant neural networks (96). Relevant for telehealth and virtual intervention delivery, other hyperscanning studies (Total N=84; N=42 dyads) observed that even when collaborating online via a cooperative multiplayer game, dyads showed elevated neural synchrony (90), indicating that phase synchronization of oscillatory activity occurs during real-time joint coordination without physical co-presence (90). Recent hyperscanning studies, even with relatively small samples (Total N=30; N=15 dyads) have found greater synchrony within dyads for a focus on the breath task synchronization as compared to a cognitive synchronization task (97). These data regions may be relevant for behavioral intervention response, due to their nature [e.g., socioemotional processing (frontal theta); learning processes (central theta)] (93).

This study is expected to add critical data that will add to the knowledge base to improve alcohol use intervention for U-EA engaged in hazardous alcohol use. Given the minimal risk to participants and greater possibility of long-term benefit to the participant as well as the greater knowledge base, the risk/benefit ratio seems reasonable. As a part of this study, all participants have the opportunity to examine their own alcohol use behavior in the context of completing related measurement instruments, and will work with a trained therapist in a group (dyadic) setting to receive an evidence-based behavioral alcohol intervention (group MI), which may help facilitate their efforts to reduce their hazardous drinking. The costs associated with participating have been minimized via the consent procedures, procedures for maintaining confidentiality, and accommodations during the fMRI hyperscanning component. The minimal costs associated with participation in this research are reasonable in relation to the anticipated benefits to the participants themselves.

### Objectives

Given their brevity, low cost, and ease of dissemination (8), the group modality is widely used to deliver alcohol intervention with youth. Even with the background research around potential peer-peer iatrogenic outcomes during group interventions with young people (98–100), we have a limited understanding of how peers, and specifically prosocial, positive peer-peer interactions may enhance intervention outcomes within the group MI platform (101). This is timely, as there has been a surge of publications within the developmental neuroscience literature around the salience and importance of peers in terms of positive neural response, behavioral risk, and prosocial choices (102).

**In sum,** we found no peer reviewed literature examining the degree to which peer-peer dyadic exchanges within the context of group intervention receipt may impact both brain and subsequent behavioral response for youth. This study will specifically examine the nature of peer-peer dyadic exchanges within group MI, and the degree to which neural synchrony during positive, prosocial peer language (peer-generated change talk) is associated with alcohol use reductions at 12 months post-intervention. This is critical to address the gap around how and why group MI may (or may not) work in this age group, to meaningfully move the needle on enhancing alcohol-related interventions and related outcomes in this high-need and underserved age group.

## METHODS

### Patient and public involvement

Not applicable.

### Trial design

A structured summary of trial design and methods is presented in Table 1. This is a <u>translational</u> within-subjects design measuring how synchrony in U-EA brain response during positive, prosocial group MI language (peer-directed change talk) is associated with behavior change at 12-months post-intervention. Within this study, two U-EAs engaged in drinking will be scheduled to attend a “Participation Day,” comprised of a baseline behavioral assessment (which they will complete alone), a single 1-hour session of group MI (completed together), and, to ensure maximum salience of the peer dyadic experience, an fMRI hyperscanning protocol on the afternoon of the same day. To determine how these factors relate to alcohol-related behavior change, youth will complete behavioral follow-up at 12-months post-intervention. Secondary outcomes will evaluate the same relationships at 3- and 6-months.

**Table 1.**
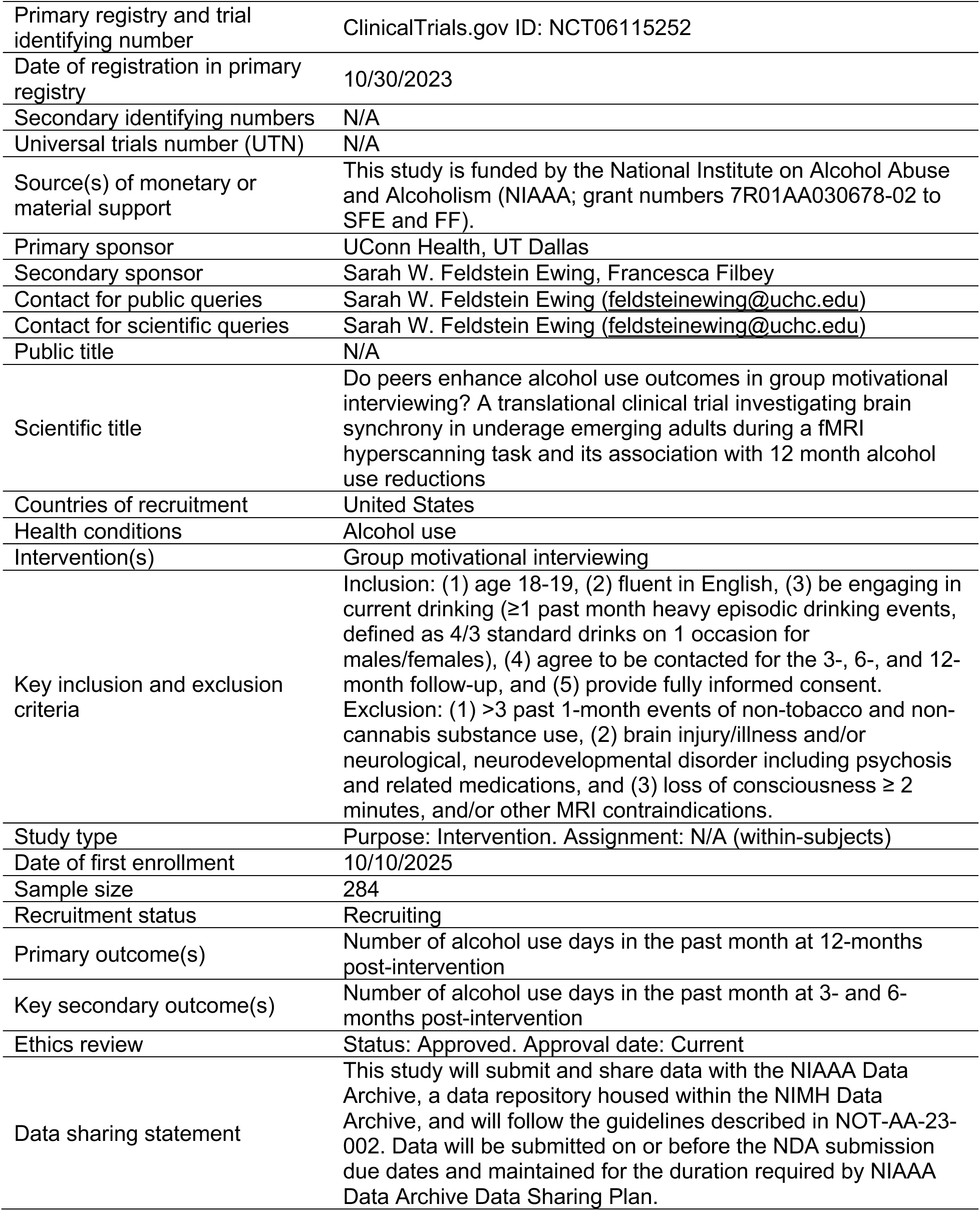
Structured summary of trial design and methods.

### Trial setting

Trial recruitment, enrollment, and study activities will take place at one university site in Dallas, TX, USA. Study interventionists provide virtually-delivered group MI from UConn Health, Farmington, CT, USA.

### Eligibility criteria

#### Participants

Inclusion criteria are purposefully broad to maximize participation and generalizability of study findings. For <u>inclusion</u>, youth must be: (**1**) age 18-19 (ensuring a sufficient follow up window to prevent youth from turning 21 during the course of participation, which could impact drinking patterns), (**2**) fluent in English, (**3**) in line with our own research and others (51, 52, 55, 103–106), U-EA will be required to be engaging in current drinking (≥1 past month heavy episodic drinking events, defined as 4/3 standard drinks on 1 occasion for males/females), (**4**) agree to be contacted for the 3-, 6-, and 12-month follow-up, and (**5**) provide fully informed consent. With respect to the fMRI component, we have attempted to maintain tight control over factors that may increase error variability and that maximize protection for our participants. Thus, our <u>exclusion</u> criteria are: (**1**) >3 past 1-month events of non-tobacco and non-cannabis substance use (e.g., methamphetamine; fentanyl), (**2**) evidence of brain injury/illness and/or neurological, neurodevelopmental disorder including psychosis and related medications (e.g., antipsychotics; neuroleptics), and (**3**) loss of consciousness ≥ 2 minutes, and/or other MRI contraindications.

#### Group MI interventionists

Study interventionists must be enrolled in or graduates of graduate-level clinical training programs (e.g., clinical psychology PhD), and have been recruited for study involvement by the UConn Health research site, under the direct supervision of MPI Feldstein Ewing.

### Intervention and comparator

#### Rationale for choice of intervention approach: Group MI

We chose group MI as the target behavioral alcohol intervention for this age group for several reasons. It has been essential to our team’s line of clinical research to develop interventions for youth that can be easily transported to and integrated within opportunistic settings (e.g., schools/universities; healthcare; juvenile justice/justice) (34, 51–54, 107–113). Operating from this perspective has allowed us to roll out interventions to non-intervention seeking youth who are at high need, such as U-EA who are engaged in drinking, but are unlikely to seek (or receive) alcohol intervention within standard care/service delivery contexts (7). Group MI has a high potential for public health reach for this age group (27, 30, 34) for several reasons, including but not limited to, its inherently cost-effective nature. While group MI is already used extensively in opportunistic care settings (8, 17, 19, 20), we still know very little about the mechanisms, especially at a neural level, for <u>how group MI operates</u>, and <u>how the role of peers may enhance youth neural and behavioral response in this intervention</u>. Because the U-EA brain is still very much in development (114) and research by our team suggests that the neural mechanisms of psychosocial intervention response varies by neurodevelopmental stage (51–55, 111, 113, 115–118), it is timely and important to specifically assess neural and behavioral response in the group MI context and within the U-EA developmental window.

#### Group MI session

In line with our previous group MI approaches to reduce alcohol and related health risk behavior with this age group, all participants will receive 1 60-minute group MI session conducted virtually by a Clinical Psychology trainee (graduate student and/or postdoctoral fellow) under the supervision of MPI Feldstein Ewing and will proceed according to our established group MI manual for U-EA alcohol use (17). The group MI session occurs in the first half of Participation Day activities, prior to the fMRI hyperscanning. This group MI aims to introduce a conversation about alcohol use and the personally experienced consequences of drinking (18, 32–34, 55, 109, 110, 119, 120). Group MI interventionists will conduct the session in an MI-consistent manner, meaning that they will be open, strength-based, affirming, non-judgmental, and empathic, with a goal of reducing resistance and highlighting ambivalence around drinking to foster and support U-EAs’ intrinsic motivation for behavior change.

Following our empirically-supported approach for MI targeting alcohol use with non-intervention-seeking youth, the group MI will start with an open-ended exploration of the dyad’s drinking behavior via eliciting the dyad’s stories about their alcohol use. Elements of the group MI intervention include, but are not limited to, an open-ended exploration of the dyad’s drinking, activities to enhance the dyad’s sense of self-efficacy, exploration of potential high-risk situations and triggers for drinking and an exploration of strategies to manage those risks and triggers, elicitation of dyadic peer-directed change talk for fMRI task, and closing of the group MI session. The ultimate goal of the group MI session is to engage the dyad in a thoughtful conversation about their drinking and the implications that their drinking may have on their lives, with an eye to bolstering and supporting the dyad’s inherent drive for behavior change. The group MI interventionist will close with a summary of the session.

During this group MI session, the group MI interventionist will track the dyadic exchange to ensure that each dyad member generates sufficient unique positive peer language statements (peer-directed change talk; voice of the peer directed toward the youth receiving the scan) for the dyadic paradigm to be extracted from the recorded group MI session and transferred to the fMRI task prior to scanning.

#### Discontinuation criteria

In line with informed consent, the group MI session and subsequent study involvement may be discontinued at any time at the request of a trial participant. The study team may withdraw an individual from the trial if the individual is disruptive, uncooperative, threatening, or physically violent towards others during a study visit.

#### Strategies to improve adherence to intervention/protocols and monitoring adherence

To ensure intervention integrity and fidelity of the intervention, all group MI sessions will follow our group MI manual (17), maintained by MPI Feldstein Ewing at UConn Health. This 60 minute group MI session was developed based on MPI Feldstein Ewing’s clinical work with youth engaged in alcohol and other substance use, and continues to be updated in line with recent publications in the MI field (17–44). In line with MPI Feldstein Ewing’s previous NIH-funded studies, she will train each group MI interventionist via the following 5 steps. Group MI interventionists will: (1) receive didactic training on the theories behind group MI, (2) be required to read the group MI study manual, (3) pass a knowledge test to evaluate their grasp of the concepts within and behind group MI (e.g., with a pass rate of at least 80% on the fidelity checklist and 100% adherence to essential therapeutic elements), (4) watch the MPI conduct a pilot group MI and discuss the details of the session, and (5) conduct two pilot group MIs, which will be observed by the MPI who will assess the group MI interventionist’s proficiency in the manualized group MI intervention. All sessions are audio-recorded for two purposes: for systematic supervision, allowing the MPI to ensure fidelity and prevent therapist drift during the weekly group MI supervision session, and for use within the fMRI scanning task.

#### Concomitant care

We do not prohibit youth from seeking/receiving any other type of intervention during the course of the trial (e.g., psychotherapy), but we do record their receipt of other intervention at baseline 3-, 6- and 12 months.

### Outcomes

Our primary outcome is past month <u>alcohol use days</u>. We are examining neural alignment (brain synchrony) in the social cognition network during positive, prosocial peer interactions (peer-directed change talk), and hypothesize that this variable will be associated with greater post-intervention behavior change (lower number of past month drinking days) at 12 months. Secondary outcomes will evaluate the same relationships at 3- and 6-month follow-up points.

### Harms

Trial harms will be passively surveilled; the trial MPIs will work closely with the study team members direcly involved in data collection (e.g., research assistants, study interventionists) to monitor for reportable events. Events will be tracked and reported to the Institutional Review Board of record in line with reporting requirements, and retained for analysis purposes as applicable.

### Participant timeline

The time schedule of enrollment, intervention, assessments, and visits is presented in Table 2. The study team will keep a rolling list of potentially eligible participants. After two U-EA have been deemed eligible and consented, our team will contact youth to ensure that there is no change in their eligibility and they will then be scheduled in dyads for “Participation Day” activities, which include the baseline assessment, the group MI session, and the fMRI scan, all conducted on the same day. The same behavioral measurement package administered at baseline will be administered via a virtual platform (121–123) for follow-up assessments at 3-, 6-and 12-months.

**Table 2.**
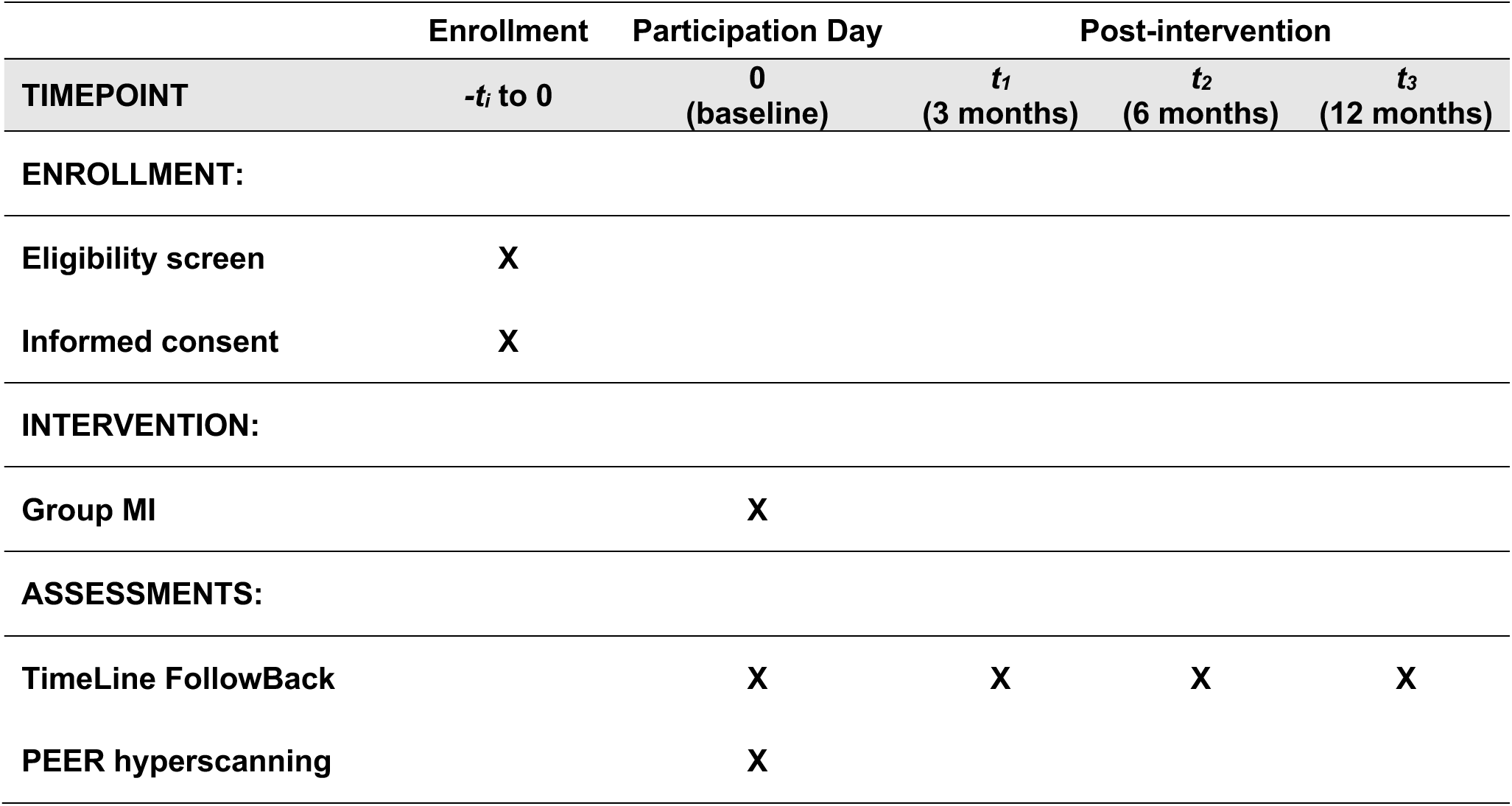
Participant timeline: Schedule of enrollment, interventions, and assessments.

|  | Enrollment | Participation Day | Post-intervention |  |  |
| --- | --- | --- | --- | --- | --- |
| TIMEPOINT | $-t_i$ to 0 | 0<br>(baseline) | $t_1$<br>(3 months) | $t_2$<br>(6 months) | $t_3$<br>(12 months) |
| **ENROLLMENT:** |  |  |  |  |  |
| Eligibility screen | X |  |  |  |  |
| Informed consent | X |  |  |  |  |
| **INTERVENTION:** |  |  |  |  |  |
| Group MI |  | X |  |  |  |
| **ASSESSMENTS:** |  |  |  |  |  |
| TimeLine FollowBack |  | X | X | X | X |
| PEER hyperscanning |  | X |  |  |  |

### Sample size

Sample size was selected to evaluate the primary research questions at a two-tailed alpha of .05 and power of .80. Estimates of effect size were conducted in G*Power 3 (124, 125). To optimize power, our a priori hypothesis was tested using regions of interest (ROIs) motivated by our previous findings. Although this is the first time we will be investigating BOLD synchrony across social cognition networks in a clinical translational study, we grounded our power based upon our previously observed associations between DMN regions and post-intervention behavior change in a similarly-aged sample (e.g., precuneus z = 32; PCC; z = 40) (51–55). Prior estimates indicated a small-to-medium sized associations (f^2^=0.07, r= -0.22) with linear multiple regression analyses requiring a minimum of 164 youth to achieve adequate power. For the associations between BOLD synchrony in social cognition regions during positive, prosocial peer-directed change talk and alcohol use reductions at 12 months, 210 subjects would be adequate to detect a medium sized effect (standardized effect size d=0.4) with 80% power. In sum, the power analysis suggests the need for N=210 youth. We project losing 15% of the proposed sample to attrition or motion, thus requiring N=248 youth.

### Recruitment

In line with previous U-EA intervention research (51, 52, 55, 103, 126), we will recruit a sample with sufficient drinking to assess mechanisms underlying behavior change within this group MI intervention. Our recruitment approach will be purposefully broad to maximize participation and generalizability to U-EA engaged in alcohol use throughout the wider community. We will utilize a combination of community-based (e.g., flyering at coffee shops, community centers, apartment complexes), campus-based (e.g., classroom sign-ups, school listservs), and social media-based (e.g., Snapchat ads) recruitment approaches that have been highly effective with this age group for our team (51, 52, 55, 127) in the broader Dallas metro area. Our team has found that this recruitment approach maximizes access, reach, and engagement of U-EAs engaged in drinking, who are unlikely to otherwise seek and/or receive alcohol intervention (128). Consistent with our prior U-EA substance use studies (51, 52, 55, 127), U-EA who observe local ads will connect with our program staff (via their call to our program line, and/or receive a phone call or text if they place their name on a sign in sheet, and/or scan a QR code) at UTD to learn more about the opportunity to participate and, if interested, be screened for eligibility.

### Randomization and blinding

As this study utilized a within-subjects design, randomization was not conducted and all participants received the same intervention. Similarly, given the focus on a single intervention, blinding to condition is not applicable to this trial.

### Data collection methods

In line with our prior studies (51–55), at the start of the Participation Day, youth will be breathalyzed using a BACtrack S80 Professional Breathalyzer to ensure BAC=0 and screened to assure the absence of fMRI contraindications (no metal in body; not pregnant as verified by urine testing for all female participants). Consistent with the MPI’s prior translational work (51–55), all participants will be required to abstain from alcohol and other substances for 24 hours prior to the scan. Only approved youth will continue with Participation Day activities.

#### Behavioral assessment

##### Primary baseline measures

Each member of the U-EA dyad will complete behavioral assessments using REDCap independently via lab laptops in separate rooms prior to their participation in the group MI. Our primary outcome is past month <u>alcohol use days</u>. We will utilize the TimeLine Follow-Back (TLFB; 129), a gold-standard calendar-based measure which provides quantity and frequency of past month <u>alcohol use days</u> in this age group.

##### Follow-up measures

We will use the same behavioral measures administered at baseline at 3, 6, and 12-months post-intervention to evaluate U-EAs’ alcohol intervention response. We will use a virtual platform for follow-up assessment (121–123). At each follow-up, youth will be emailed unique links to complete validated self-report measures via a secure online data collection system (REDCap) (130), and will complete a brief online or telephone interview with project staff.

#### fMRI (PEER) hyperscanning session

The fMRI hyperscanning session will be the last activity of Participation Day. Between the group MI and scan session, study staff will extract the dyadic peer-directed change and sustain talk generated during the group MI session and integrate them into the dyad’s fMRI task. The fMRI data will be collected using two 3 Tesla Siemens Prisma scanners at the Brain Performance Institute at the University of Texas at Dallas. Images will acquired using a T2*-weighted echo-planar imaging (EPI) sequence with the following parameters: TR/TE=800/37ms, flip angle=52°, FOV=208x100mm (ROxPE), matrix=104x100(ROxPE), slice thickness=2.0mm, 72 slices; 2.0mm isotropic voxels, multiband factor=8, echo spacing=0.58ms, BW=2290 Hz/Px. 3. High-resolution structural images will be obtained using a T1-weighted sequence for anatomical localization and normalization.

##### fMRI (PEER) hyperscanning task (Figure 1)

Both participants in a dyad will be scanned simultaneously in separate but identical scanners separated by a shared control room. Real-time audio and visual stimuli will allow for a naturalistic interaction. Task events will be synchronized between the two scanners using specialized software to ensure precise temporal alignment of neural and behavioral data.

**FIGURE 1.**
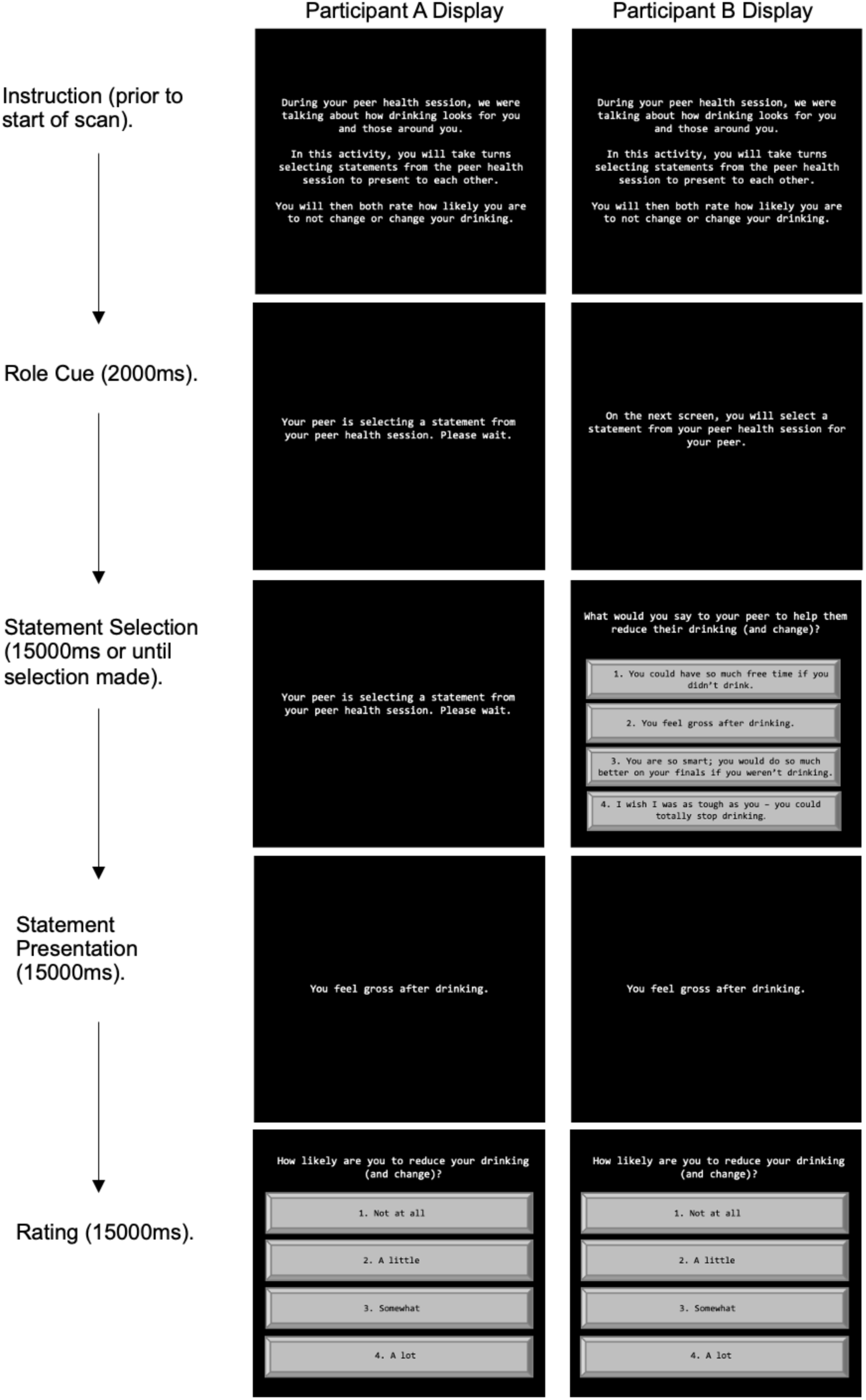
PEER hyperscanning task: The example below walks through the visual presentation during peer-directed change talk condition, with each column showing the simultaneous presentation of identical or different displays based on trial stage and participant role.

MPIs Feldstein Ewing and Filbey have pioneered the examination of within-session factors (client language; therapist language) in the youth brain (51–55). For this study, we will build directly upon our validated Neural In-Session Language (NILE) Task used in our previous NIAAA-funded translational youth R01 (R01AA023658; project dates 4/17/15-3/31/22) and published in NeuroImage:Clinical (55). Our prior study indicated that salient within-session factors during individual-level interventions (there, therapist language) influence the developing brain and that brain response was significantly associated with youth intervention outcomes (51–55). However, different from that study, we will shift the focus from looking at youth response to therapist:youth exchanges within an individual-level intervention (individual MI) to the current study’s novel context of peer:peer exchanges in a tandem, hyperscanning fMRI approach and in a group-level intervention (group MI).

In the PEER hyperscanning task, U-EAs will be presented with peer-directed positive, prosocial statements (peer-directed change talk), generated during their group MI session. Consistent with our previous studies, we will present each dyad with a version of the task that has been specifically articulated to the clinical exchanges directly extracted from their group MI session. The PEER hyperscanning task begins with an Instructions screen, with accompanying pre-recorded audio of the group MI interventionist, to orient the dyad to the task. During the task, participants will simultaneously be presented with audiovisual presentations of 8 talk statements (4 peer-directed change statements and 4 peer-directed sustain statements) recorded during their peer health session in pseudorandom order. At the start of each trial a participant will be chosen randomly and will select a statement to present to the other from the following conditions: Peer 1 Select - peer-directed change statements (4 trials), Peer 1 Select – peer-directed sustain statements (4 trials), Peer 2 Select – peer-directed change statements (4 trials), Peer 2 Select – peer-directed sustain statements (4 trials). Each statement type will be presented four times per participant, resulting in a total of 16 trials. Each trial begins with a Role Cue screen that will designate which participant will make the statement selection, and which will wait while the other makes the selection. This will be followed by a Statement Selection phase where audiovisual stimuli (in the interventionist’s voice) will be presented asking the designated participant to select a statement from a list of four statements using the button box, while the other participant waits. Next, the participant’s selection will be shown via audiovisual stimuli during the Selection Presentation phase where the selected statement will be delivered visually on the screen in addition to audio recording of the participant (recorded during the health session). Each trial will end with a Rating phase where both participants will be asked to rate how likely they are to change their drinking behavior via 1 – 4 rating scale (corresponding to a four-button button box) read in the group MI interventionist’s voice. There may be a jitter between trials during which a fixation cross will be presented.

#### MI session coding fidelity/reliability

PI Feldstein Ewing has been coding client language in MI since 2004 and culling client language for fMRI tasks since 2007. As with PI Feldstein Ewing’s prior NIH-funded studies (51–55), in order to learn how to identify statements for the task, study staff will be required to read the study manual and conduct 5 practice coding sessions by identifying and extracting 4 types of statements in mock interventions conducted by the study team – including in terms of having statements that are sufficiently meaningful (of a sufficient intensity) of the correct valence. This protocol has been highly effective in training study staff to successfully identify and extract statements directly from the recorded MI session and transfer these statements to the fMRI task scan (51–55).

#### Retention

Youth engaged in alcohol use are a difficult population to track. Yet, with intensive tracking procedures, and predominantly all online follow-up assessment measures (such as proposed here), our success in retaining a similar age group of substance-using U-EA (∼90% of youth at the most distal follow-up) suggests that we successfully can retain a majority of participants by implementing those strategies here. As examples, our study team is trained in techniques to build and maintain rapport, given resources to accommodate U-EA contact method preferences (e.g., study-dedicated phones with texting capability), and provided with MPI support to troubleshoot participant contacting and scheduling difficulties, as MPI Feldstein Ewing is an expert in the recruitment and retention of underrepresented youth, and has published extensively on how to retain substance using and other high risk youth in longitudinal research (131–133). In addition, the trial design allows for fully remote follow-up data collection (using a combination of REDCap and telephone), eliminating the burden of in-person visits beyond the baseline Participation Day. In the infrequent instance an enrolled participant declines further participation, study team members are trained to honor the participant’s autonomy while respectfully collecting data regarding discontinuation reason(s), to ensure a thorough CONSORT post-trial and adequate data for analyses, as applicable.

### Data management

Behavioral assessments will be captured electronically via secure online platform, REDCap. All data will be managed on a UTD secure server only accessible by approved study personnel. All data management and manipulation will be conducted via widely utilized software packages including, but not limited to, AFNI, R, and Matlab. It will be possible to share de-identified data in several formats.

#### Data quality and control

Standard Operating Procedures (SOPs) for the study protocol will be developed. All RAs will be trained to ensure rigorous data collection. Data assurances will be performed by the RAs for missing data or implausible responses. The RedCap Data Quality module will be utilized to identify data discrepancies and other data issues. Additionally, data validation and spot checking will be conducted by a supervisor to check for data completeness and internal consistency of responses. The REDCap database will maintain an audit trail with time-date stamps of data entry and all changes that are made to the data.

#### Data security

Every effort will be made to ensure data security. Any paper materials will be double-locked – i.e., stored in locked file cabinets in locked offices that belong to the study team. Electronic data, such as REDcap and MRI, will be kept secure with encryption and password protection. Data will be de-identified and labeled with a random number identification code that does not contain personal identifying information.

#### Data storage

Electronic data will be stored using UTD-approved platforms (Box, REDCap). Consent forms and data collected on paper will be stored and double-locked in a secure archive for long term storage. All paper source records will be retained for a minimum of 10 years from the point of publication of data on the primary outcome. Electronic data will be stored for a minimum of 10 years following study completion, with regular checks to make sure that the data are still viable.

### Statistical methods

#### Data processing

Functional and anatomical MRIs will be processed and analyzed using the Analysis of Functional NeuroImages (AFNI) software package (https://afni.nimh.nih.gov/afni/ version AFNI_22.0.03). The T1-weighted anatomical images will be semi-manually segmented producing a skull-stripped image. A secondary segmentation and warping will then be applied using @SSwarper to the skull-stripped image to refine the segmentation and warp the image to conform to the Montreal Neurological Institute stereotactic space (MNI152 2009). We will generate the functional processing pipeline using AFNI’s afni_proc.py script that discards the first 6 EPI volumes, implements slice timing and field map distortion corrections, registers fMRI volumes to the minimum outlier, aligns and warps volumes to the template space provided by the anatomical transformation, spatially smooths using a Gaussian filter (5 mm full-width-at-half-maximum), and mean scales voxels time series to 100. Frame-to-frame displacement will be calculated for each volume.

##### First-level univariate models

Model fitting will be computed with the 3dDeconvolve and SPMG2 basis functions, as well as 3dREMLfit estimation of auto-correlation. Stimulus timing files from the fMRI task will be entered into models as well as nuisance regressors estimating frame-to-frame translational and rotational motion and signal intensity outliers.

ROIs will be defined using the probabilistic Harvard-Oxford cortical and subcortical structural atlases (distributed with FSL; https://fsl.fmrib.ox.ac.uk/fsl/fslwiki/Atlases). ROIs will be constructed as 5mm radius spheres.

#### Primary outcome analysis

We will leverage the dynamic nature of the PEER hyperscanning task to examine neural responses in the U-EAs social cognition network (medial prefrontal cortex [mPFC], superior temporal sulcus [STS], temporoparietal junction [TPJ]) as participants engage with a novel peer from their group MI session. We will examine the degree to which synchrony of BOLD response that occurs while hearing and seeing positive, prosocial peer-directed health promotive language (peer-directed change talk) directly generated during and extracted from the group MI session is associated with youth behavior change (past month drinking days) at 12 months. It is expected that the BOLD synchrony during peer-directed change language will be associated with behavior change at 12 months. Primary outcomes include fMRI-based hyperscanning metrics (synchrony of BOLD response in social cognition networks) and behavioral measures (Timeline Followback; TLFB) to assess past month day drinking at 12-months. Secondary outcomes will evaluate the same relationship at 3- and 6-months.

Inter-brain synchrony will be quantified to capture moment-to-moment coordinated neural responses between each dyad during the hyperscanning fMRI task. Mean BOLD time series will be obtained for each ROI and temporally aligned across participants. Inter-brain synchrony will be quantified using Pearson correlation coefficients (r) between the BOLD time series of dyad members within ROIs encompassing the U-EA social cognition network (mPFC, STS, TPJ). To capture moment-to-moment neural alignment, correlations will be computed using a sliding-window approach, with window size and step size selected to balance temporal resolution and signal stability. Correlation values will be Fisher Z-transformed prior to group-level analyses to ensure normality.

These synchrony metrics (i.e., ROI z-scores) will be used to test our hypothesis that greater neural alignment in the social cognition network during positive, prosocial peer-directed change language will be associated with greater behavior change. Permutation testing and multiple comparison correction will be applied when assessing statistical significance.

#### Missing data

Co-I Yang’s is expert in analyzing longitudinal data and developing methods to handle both missing-at-random and missing-not-at-random data in longitudinal randomized trials (134, 135).

### Data monitoring

#### Data monitoring committee

This trial does not require a Data and Safety Monitoring Board, as it does not incorporate components that may warrant additional monitoring and review beyond MPI, IRB, and funder oversight (e.g., multiple clinical sites, blinding, high-risk interventions, vulnerable populations), and it is not a Phase III clinical trial. Interim analyses and stopping guidelines are not part of the trial protocol due to the trial design and low risk status.

#### Trial monitoring

MPIs will self-monitor study site recruitment, enrollment, and consenting procedures; trial intervention delivery and fidelity; event tracking and reporting; and data collection and management procedures to ensure trial conduct is in line with the trial protocol and Research Ethics Approval.

## ETHICS

### Research ethics approval

This study has human participant Research Ethics Approval through University of Texas at Dallas. Per the IRB Reliance Agreement in place for this trial, UConn Health will rely on UTD as the IRB of record.

### Protocol amendments

All members of the UTD and UConn Health study team meet at least once per month at a standing virtual meeting, with the opportunity to discuss the need for protocol modifications. If the MPIs determine that a protocol amendment is necessary, the study team member at the applicable site submits the materials to the ethics oversight board. Protocol modifications are not implemented until the relevant ethics board has reviewed and approved the submission.

### Consent or assent

As detailed in *Recruitment*, IRB-approved study team members with current human subjects protections training describe study content and procedures to interested U-EA via phone. The staff members are explicit that participation is completely voluntary, participants can opt out of the trial at any time without repercussion, and skip data collection questions that they do not feel comfortable answering. Specific eligibility criteria are not shared with participants to further protect participant privacy. If an interested U-EA is eligible to participate, they will complete consent forms on paper at the start of their scheduled Participation Day. This system allows the participant to review the entire consent form in person, discuss the study in detail and have questions answered with a UTD study team member, as well as allows the UTD study team member to ensure the potential participant understands the consent form before choosing whether or not to complete the form. The trial consent form specifies that participant data may be used for future research and details the conditions under which various data types may be used. This written informed consent will be obtained prior to engaging in any trial activities; individuals who decline to participate will be thanked for their time and interest and will not be enrolled in the trial.

### Confidentiality

All collected data will be coded using a subject ID number that is not derived from participant identifying information, and only trained UTD research assistants and MPI Filbey will have access to the password-protected master subject identification sheet linking contact information with subject ID numbers. This linking file will be destroyed at the conclusion of the study when participant contact information is no longer necessary for data collection and retention purposes. Consent materials will be stored electronically on encrypted computers in locked offices, and paper documents in locked cabinets in locked UTD study offices. Consent files and participant contact information are kept separate from participant study materials.

The people who will have access to trial data include members of the research team. All study staff complete required institution-specific training modules for clinical trials research with human subjects, and are trained on lab and study-specific protocols to protect participant confidentiality. Identifiable data will not be shared with investigators outside of the research team without an executed Data Use Agreement.

### Ancillary and post-trial care

This is a minimal risk trial and the study team has taken great care to reduce or, when possible, eliminate possible risks of harm to trial participants. As a result, this trial does not include plans for ancillary and post-trial care.

## Notes

**Primary funding:** This work was supported by the National Institute on Alcohol Abuse and Alcoholism (NIAAA; grant numbers 7R01AA030678-02 to SFE and FF). NIAAA had/will have no role in the design, conduct, analysis, and reporting of the trial, including any authority over these activities.

### Competing Interest Statement

The authors have declared no competing interest.

### Clinical Trial

https://ClinicalTrials.gov ID: NCT06115252
ID: NCT06115252

### Author Declarations

IRB of University of Texas at Dallas gave ethical approval for this work. IRB of UConn Health will rely on UTD as the IRB of record per IRB Reliance Agreement.

